# Improving HIV Outcomes in Miami’s Black populations with clinic-based community health workers protocol: The integrated navigation and support for treatment adherence, counseling, and research (INSTACARE) randomized controlled trial

**DOI:** 10.1101/2024.12.11.24318859

**Authors:** Sonjia Kenya, BreAnne Young, Sebastian Escarfuller, Deborah Jones-Weiss, Olveen Carrasquillo, Ana Riccio, Pan Yue

## Abstract

Miami-Dade is an HIV epicenter where Black populations experience excess AIDS-related deaths due to poor medication adherence, which prevents achieving an undetectable HIV viral load (VL). A promising approach to improving HIV outcomes in Black populations has been the use of community health workers (CHWs). Evidence shows CHWs trained in motivational interviewing (MI) may further improve outcomes, however little data exists about Black CHWs trained in MI who support Black patients with HIV. While CHWs traditionally help address social determinants of health in nonclinical locations, there is less information on CHWs who provide support in clinical settings, which may result in even greater improvements in HIV outcomes. To examine effects of CHWs trained in MI who provide HIV care in both clinic and community settings, a randomized controlled trial (RCT) is being conducted in Miami-Dade’s largest public hospital. The Integrated Navigation and Support for Treatment, Adherence, Counseling, and Research intervention is a RCT of 300 Black adults with an unmanaged HIV VL (> 200 copies/mL). CHWs trained in MI are embedded into HIV clinical care teams and participate in hospital rounds with clinicians treating inpatients. Participants randomized into the CHW intervention arm receive 12 months of CHW-led health education and assistance with health care navigation and social services. The primary outcome is change in HIV viral load suppression at 12 months. Secondary outcomes include changes in medication adherence and social determinants of health. Study enrollment began in 2023 and will be completed by 2027. The first results are expected to be submitted for publication in 2025. INSTACARE is one of the first RCTs to examine effects of clinic-based support provided by CHWs trained in MI on Black populations with an unmanaged VL and will provide evidence on the impact of such strategies on medication adherence and social determinants of health.

## INTRODUCTION

Miami-Dade is an HIV epicenter where Black adults account for 15% of the County’s population and 64% of AIDS-related deaths [1,2]. Despite availability of proven HIV treatment and prevention resources, the HIV incidence rate in Miami persists as one of the highest in the nation and the region’s Black communities experience some of the greatest HIV disparities [3,4]. Locally, 1 in every 38 Black adults has been diagnosed with HIV compared to 1 in 136 White adults and 1 in 129 Latin adults [1]. Optimal HIV health is indicated by viral load (VL) suppression, achieved only through antiretroviral therapy (ART) adherence and care continuity. Coordination of health care delivery among various providers, VL monitoring, and adherence to non-HIV medications among those with comorbidities, make HIV a difficult chronic disease to manage. Disproportionately poor social determinants of health (SDOH) in Miami’s Black communities further compound these challenges. For these reasons, Black people living with HIV (BPH) in Miami suffer the County’s lowest rates of care engagement, retention, and viral load suppression.

The causes for this lack of engagement in HIV care are complex and influenced by a multilevel interaction of disease, behavior, and SDOH, including poverty, homelessness, unemployment, and stigma [5–8]. Miami’s Black population accounts for nearly half of the poor, over 40% of the homeless, and excess unemployment (15%) that nearly triples that of whites [9,10]. HIV stigma further exasperates these challenges, increases the risk for ART non-adherence, and contributes to BPH falling out of care [11–13].

To date, one of the most promising interventions to improve care continuity among BPH involves using community health worker (CHW) strategies [14–16]. CHWs are culturally-congruent lay health workers who serve as an interface between the community and the health care system in highly vulnerable populations [17–19]. They provide health education in culturally and linguistically appropriate ways, help patients access effective health care by functioning as patient navigators, and assist with nonmedical barriers such as social issues [20–22]. Our work, along with many others’, has shown CHWs effectively address SDOH, leading to improved care engagement, HIV outcomes, and viral suppression among BPH [14–16].

Additionally, research suggests CHW interventions could benefit further from the use of theory- driven intervention strategies, such as Motivational Interviewing (MI), especially when working with Black populations. While MI has traditionally been used by health practitioners to influence behavior change among their patient populations, in recent years, it is increasingly employed by CHWs working with vulnerable populations, however less information is available about the impact of CHW-led MI among Black populations with poorly controlled HIV [23]. As members of the community, CHWs are able to successfully use MI to explore and evoke change talk that elicits a patient’s intrinsic desire, ability, and reasons to make a behavioral change [23].

CHWs traditionally provide patient support in nonclinical locations, including homes, recreation centers, or local parks [20,21]. Although a wide body of research demonstrates the clinical value of community-based CHW support addressing SDOH and HIV outcomes, less research exists on the impact of CHWs who also provide support in clinical settings, which may result in even greater improvements in HIV outcomes.

To examine the impact of CHWs who provide HIV care in both clinic and community settings, this research team developed the **Integrated Navigation and Support for Treatment, Adherence, Counseling, and Research (INSTACARE) intervention** to address the SDOH underlying poor HIV outcomes among BPH in Miami, FL.

## METHODS

### Preliminary Research

INSTACARE builds upon the findings of a formative pilot study [24,25] which aimed to 1) better understand how clinic-based CHWs embedded in South Florida’s largest safety-net hospital could contribute to improved HIV outcomes in BPH, and 2) the feasibility of this approach. This preliminary research was conducted in two parts. Phase 1 involved interviews with patients, caregivers, and clinic staff to assess perceptions on the barriers and facilitators to HIV care [24].

Results suggested integrating CHWs into HIV clinical teams could be an efficacious approach to overcome systemic barriers to HIV care and address unmet social needs that are beyond a physicians’ purview. Using this feedback, Phase 2 included the development and implementation of a 3-month clinic-embedded CHW intervention to improve ART adherence among BPH in Miami-Dade County [25]. Among 10 participants, this study found CHWs were a practical and feasible enhancement to HIV clinical care teams. Further, CHWs’ ability to provide targeted social and clinical support suggested this model could be a promising strategy to achieve sustained viral suppression and care engagement for BPH.

### Conceptual Design

The Integrated Navigation Services for Treatment Adherence, Counseling, and Research (INSTACARE) study (R01MD08187) is a two-arm 1:1 randomized control trial (RCT) examining the effects of 12-months of CHW support on HIV viral load (VL) among 300 Black people living with HIV in South Florida. INSTACARE is grounded in the Social Ecological Model, a multidimensional theoretical framework that broadly examines how the interactions between one’s social conditions and environment influence health behavior [26]. The focus of this design emphasizes CHW intervention models as a feasible strategy to address systems level barriers to care across multiple domains, including individual, interpersonal, organizational, and community. The study is registered with ClinicalTrials.gov (NCT04663152) and ethical approval was obtained by the University of Miami Institutional Review Board (IRB #20201234).

### Study Setting

INSTACARE is based in Miami-Dade County, which has a culturally and ethnically diverse Black community unlike anywhere else in the Nation. Home to the largest enclave of Black immigrants in the U.S., one-third of Miami’s Black residents are foreign-born, many from Caribbean or South American descent [27]. While composing less than 20% of the County’s residents, this population represents 64% of AIDS-related deaths [28].

Participants for this study are being recruited from South Florida’s largest public health system; a network of hospitals, specialty centers, urgent care facilities, and community clinics, including the County’s safety-net hospital, Jackson Memorial (JMH) [24]. As the teaching hospital for University of Miami Miller School of Medicine (UM), JMH and UM share a campus in Downtown Miami. JMH is staffed by an interdisciplinary treatment team of hospital clinicians, including UM faculty. In 2022, JMH providers ascertained over 750 of their patients living with HIV had not achieved viral suppression, the majority of which identified as Black [29]. Participants were recruited from this patient population, many of whom face significant barriers to care, such as extreme poverty, homelessness, substance use, mental illness, domestic violence and other social challenges associated with suboptimal medication adherence.

### Inclusion and Exclusion Criteria

Eligible participants are Miami-Dade residents aged 18 years or older, who self-identify as Black or African American, and have unmanaged HIV, as indicated by a VL > 200 copies/mL. Initially, eligible participants (referred to as clients by the study team) were expected to have a detectable VL at the time of study enrollment. However, three-months after enrollment began, the investigators realized that numerous participants referred by HIV clinicians often had undetectable viral loads during study screening, despite clinical documentation of an extensive history of unmanaged HIV. This trend prompted investigators to acknowledge that recent advancements in HIV medications can help PLH achieve viral suppression in as little as one to three weeks. Thus, eligibility criteria were expanded to capture BPH who have a history of unmanaged VL. Specifically, participants are eligible for enrollment if they present documentation of an unmanaged VL (> 200 copies per mL) anytime within the six-months prior to enrollment.

### Recruitment

Recruitment for this five-year study began April 2023. To date, most participants have been recruited from the HIV treatment clinic within JMH. Prior to enrollment, the CHW team developed a repertoire with the clinical staff to facilitate easy integration into the clinical care system. This included an in-service training for the clinical staff conducted by the research team in March 2023 to provide information on the study aims, target population, and eligibility criteria. Study brochures were placed in clinic waiting areas, replenished weekly, and distributed to all clinic healthcare providers and case managers to increase the study’s visibility. Providers and case workers also shared these brochures with potentially eligible patients. Additionally, CHWs attended weekly in-patient rounds in the Infectious Disease Ward and consulted with the clinical team case managers to identify potentially eligible participants.

### Consent and Enrollment

In this ongoing study, potential participants are often introduced to a CHW by HIV providers and case managers during their clinic visits. Patients who express interest are connected to a study team member who provides a detailed description of the study and verifies the patient’s eligibility criteria, including viral load >200 copies/mL. Qualifying patients are then linked to a CHW to schedule an intake appointment, at which time informed consent is obtained and the baseline assessment is administered.

Informed consent (IFC) and all study data are captured electronically via the University’s electronic data management system, REDCap. Participants are provided a paper copy of the informed consent for their records and CHWs read through the entire document in the participants preferred language. Study materials, including informed consent forms, are translated to Spanish and Kreyol for the patient’s comfort. Participants are asked to digitally sign the IFC; participants unable to sign the digital consent are provided a paper copy to sign for our records. The baseline assessment is then administered orally, and participant responses are recorded to REDCap by the CHW. The enrollment process takes approximately 1.5 – 2 hours to complete and participants are compensated $125 for their time.

Enrollment is frequently conducted at the INSTACARE offices located on the UM-JMH campus but can also be performed off-campus at a mutually agreed public location. CHWs are encouraged to use the buddy system, when possible, for field-based enrollment. Staff electronic devices (including mobile devices and tablets) are equipped with cellular hotspots and unlimited data to facilitate electronic data collection, and CHWs are trained to identify locations with ample cell service when feasible. Participants retained throughout the study are compensated $400 for their time participating; $125 at baseline and exit, and $50 at the 3-, 6-, and 9-month checkpoints.

### Randomization

Following the IFC and baseline assessment, participants are assigned to the CHW intervention or usual care (control) group using stratified block randomization. The sample was stratified by sex (male, female) with a fixed block size of six and a 1:1 randomization allocation. While the principal investigator and program coordinator are blinded to study allocation, both participants and CHWs are aware of the group to which each participant is randomized.

### Study Procedures

#### Community Health Worker Intervention

##### CHW Characteristics

CHWs employed during the pilot study were subsequently hired for this intervention as well. Building upon the pilot framework, additional CHWs hired for this study were culturally congruent with the population of focus, highly familiar with local Black communities, and had prior experience in service delivery (e.g., social work, HIV testing, linkage to care support, education services, etc.) among Black populations. The CHW team is also multilingual, with multiple members speaking Haitian Creole and Spanish. CHWs are assigned a caseload of 10- 15 intervention clients, depending on the complexity of the client. Complexity refers to the difficulty of a client’s care needs, ranked on a three-point scale from low to high. The complexity of each case is determined by the CHW and is subject to change throughout their enrollment period.

### CHW Training

All CHWs complete HIV pre- and post-counseling coursework through the Miami-Dade Florida Department of Health [31]. As shown in Table 1, this blended learning series includes two online courses and an in-person classroom training centered on counseling, testing and linkage to care. Each online component takes approximately five hours to complete and covers the history, demographics, and transmission of HIV/AIDS, strategies for counseling on prevention and treatment, and HIV test administration. The in-person training was a four-hour workshop providing hands on learning through open discourse and roleplay activities.

CHWs also receive an introduction to Motivational Interviewing theory and practice tailored specifically for HIV healthcare workers. The four session series provides a didactic overview of the MI process and its practice through demonstration practices, guided roleplay, and skills evaluation by self and MI instructors.

Additionally, CHWs complete all University of Miami required training modules for human subjects’ research, including coursework through the Collaborative Institutional Training Initiative (CITI), which covered human subject protections, HIPAA guidelines and policies, ethical considerations for vulnerable populations, and current regulatory and guidance information. CHWs also receive ongoing training including continuing education modules on topics such as motivational interviewing, clinic and insurance navigation, and social support resource navigation. Weekly meetings are also held with a CHW supervisor to obtain performance feedback and discuss individual cases.

### Motivational Interviewing

INSTACARE CHWs are also trained in Motivational Interviewing (MI), a patient-centered interview technique intended to elicit behavior change by increasing intrinsic motivation [31]. Historically, MI has been used by trained clinicians and counselors to support patients with substance use and addiction disorders [32,33]. Conversations are held in such a way that the participant should increase their motivation to make changes based on the values and beliefs they hold. Immigrant communities struggling with the unfamiliarity of the health-care system, limited transportation, and fear of deportation, may benefit from MI as the conversations seek to address potential barriers in depth with the CHW [31]. For example, the depth of the MI conversation may lead a CHW to support a participant in identifying strategies to minimize deportation risk, such as by relying on a friend or family member to help with transportation [31].

### Months 1-3: Individualized Activation

The CHW intervention is divided into 3 phases: activation; commitment; and maintenance (Table 2). The activation phase takes place during the first three months of the intervention. Within one week of enrollment, clients are assigned a CHW, and a meeting is scheduled for a comprehensive, face-to-face evaluation of the client’s social and environmental conditions. This evaluation includes include a discussion of (1) the client’s socioeconomic experience, such as housing and income stability or food availability; (2) potential barriers within their built environment and access to reliable transportation; (3) the presence of social support systems; (4) experiences with the criminal justice or immigration systems; and (5) knowledge of the healthcare system, including ability to communicate with providers, familiarity with their healthcare coverage, understanding their HIV status and treatment plan, and general system navigation. CHWs also discuss current lifestyle behaviors, including drugs/alcohol use and sexual risk behaviors. While INSTACARE is focused on improved VL through increased ART adherence, CHWs also address these issues from an overall health and general well-being approach.

For the first three-months, CHWs engage clients weekly in problem-solving processes to set priorities for immediate problem resolution. Personal health goals are established for VL control, such as developing strategies to improve ART adherence, addressing barriers to care, and resolving ambivalence about HIV treatment. A plan is developed to accomplish those goals and review results. CHWs also accompany clients to their medical appointments and review treatment plans to ensure they understand provider recommendations.

### Months 4-6: Strengthening Commitment to Manage HIV

Months 4-6 focus on practicing new behaviors including building self-efficacy and focusing on long-term change benefits. Depending on each client’s development, in-person meetings will gradually be replaced with phone-based support as they improve skills to manage HIV. Clients also continue to review their personal health goals regarding VL control and revise their plan to accomplish those goals as needed. By Month-5, contacts outside of clinic appointments or other support services are largely be made by phone as clients approach the maintenance phase.

### Months 7-12: Maintenance

After the first six months, participants will enter the Maintenance Phase, aimed at continued commitment to sustaining behavior change. Clients are contacted two to three times per month to check on their status. These contacts may also include appointment reminders, facilitating contact with providers, and rescheduling appointments as needed.

Findings from an earlier RCT among this demographic found mean viral load among participants in both the control and intervention groups began to increase, or “rebound,” after 9 months, suggesting a high-level of CHW activities may be needed to maintain the improvements achieved during earlier months [34]. Accordingly, the INSTACARE CHW protocol highlights this increased risk of rebounding. Specifically, CHW training emphasizes the importance of checking client progress and facilitating additional support services 90 to 120 days before study exit.

Such cases are discussed with the team of investigators to determine the appropriate case-by-case response, such as modifying service intensity or altering the type of support delivered (i.e., review ART adherence strategies or verify client efficacy to maintain and social services, including food stamps, housing assistance, transportation vouchers, health insurance, and mental health support). As the 12-month enrollment period nears conclusion, clients should be near independence and feel comfortable managing their HIV care autonomously. Final CHW activities may include linking participants to additional community- and clinic-based resources for ongoing social support beyond their study participation period.

### Control Group: Enhanced Usual Care (EUC)

Patients randomized to the control arm receive their usual HIV services according to their standard care plan, as well as HIV management brochures, and a check-in call from the study team every three months to remind them to get their VL assessments completed each quarter. Services may vary based on the needs identified by each patient’s care team, as well as their insurance policy. For example, some patients are assigned HIV case workers, members of the clinical team that assist patients with medical follow-up, socioeconomic support, and other outside referrals. INSTACARE CHWs work with the case managers to ensure patients in the control arm are reminded of their clinic appointments and have support for care continuity.

### Data Management

Physical and electronic data are stored at the INSTACARE Research Office based at the University of Miami Miller School of Medicine. Electronic data is stored on the University’s Research Electronic Data Capture (REDCap) system, a cloud-based Web app for on- and offline data capture. This data management system is managed by the University and all data collected is backed up to institutional servers daily. Staff are provided with University-issued computers, tablets, and mobile devices equipped with password-protection and University-issued encryption. Data are reviewed by the data manager on a biweekly basis for missing values or potential inconsistencies. As needed, these discrepancies are reviewed with the study team and statistician.

### Sample Size & Statistical Power

Sample size was determined by analyzing the VL suppression data from the pilot study in which participants enrolled in the intervention achieved a significant VL reduction of 1.5log10 copies/mL versus the usual care arm. Based on this analysis, the proportion of patients achieving viral suppression in the CHW arm was estimated to be at least twenty percentage lower than those randomized to the control group. Using an alpha significance level of 0.05, two-sided t-test analyses estimate that 147 participants per study arm are needed to detect the minimum difference between groups with 90% power. Thus, a sample size of 300 participants over the course of the 48-month recruitment period was selected.

## RESULTS

At the time of this writing, 95 BPH have been enrolled into INSTACARE, 42 of whom have been randomized to the intervention The first participant was enrolled in July 2023 and enrollment is projected to end early 2026. A manuscript describing the baseline characteristics of the full sample is expected to be completed and submitted for publication in 2027.

### Primary Outcome

The primary outcome is a change in viral suppression from baseline to exit at 12-months. Based on the outcomes of our previous studies, participants admitted to the EUC group are expected to have low rates of viral suppression, and participants within the intervention arm are estimated to have least a 20% higher proportion of viral suppression compared to the control arm. With a sample size of 300 virally unsuppressed BLH randomized 1:1, and an assumption 25% viral suppression in the control group, this study will have above 91% power to detect at least a 20% absolute difference between the two study arms.

### Secondary Outcomes

To measure the impact of CHW support, the mode of service delivery (e.g., face-to-face interactions, phone calls, text messages) and the setting in which services occurred (e.g., clinic, home, virtual, community setting) are tracked for each patient-CHW interaction. Social support services provided by the CHWs (e.g., health education, health system navigation, linkage to social services) were also captured using a detailed log system for CHW field notes.

Additionally, as shown in Table 3, several measures were used to capture the influence of CHWs on socioeconomic and psychosocial outcomes across multiple domains at baseline, 6- month follow-up, and study exit. The entire survey took approximately 45-60 minutes to complete and examined how the intervention may have improved client outcomes, including health-related social needs, quality of life, and self-efficacy. For these secondary outcomes, test statistic used is the two-sided Z-Test with unpooled variance and .05 significance level; there will be over 80% power to uncover a standardized difference of .10 between groups.

### Analytic Plan

Using a logistic GEE model, a type III contrast between CHW and UC arms will be estimated in a model with a binomial distribution and logit link for the primary outcome. The GEE model will include both 12-month HIV viral suppression status and HIV viral suppression status at 3-month intervals. Univariate and multivariate models will be used to calculate odds ratios and corresponding 95% confidence intervals for associations between viral suppression at 12- months and the two study arms, controlling for covariates such as sociodemographics and potential confounders. In addition, stratified analysis will be examined for gender, ethnicity, and immigration status differences. Two-way interaction terms between various study variables and the study arms may also be included in the models. Standard diagnostic tools will be used to assess model fit, If needed, alternative analyses may explore others model to account for such missing data such as last valued carried forward or multiple imputation.

## DISCUSSION

This study is designed to examine the impact of community health workers on HIV viral load suppression among Black adults with a history of treatment nonadherence in Miami-Dade County, a high priority area designated for intervention by the U.S. Government’s Strategy to End the HIV Epidemic. Among this initial sample, a substantial proportion face significant barriers stemming from their social determinants of health. More than half are unhoused or have unstable housing, and over 75% have a difficult time paying for basics, such as food or rent. By addressing the underlying socioeconomic barriers this population faces, INSTACARE seeks to mitigate the social determinants impacting HIV outcomes for PLH. This approach is further enhanced by the use of Motivational Interviewing. As a strategy principled on meeting people where they are [23], MI is well suited to address the unique barriers impacting ART adherence among individuals living with HIV [35]. While research on the impact of CHW-led motivational interviewing is still emerging, preliminary trends among this sample population show promising results for patients the receive with MI-trained CHW support.

### Limitations

Participants lost to follow up are a major expected challenge among this patient population and prior HIV research reports attrition rates between 15% - 35% [36,37]. To limit attrition, particularly in the comparison group, participants are asked during intake to provide the contact information for two persons to contact if we are unable to reach them. Updated contact information is also requested from all participants every three months to maintain open lines of communication. Missing data is also a pervasive problem in HIV behavioral research, primarily due to participant drop out [38,39]. During the analytic phase, missing data will be carefully assessed for potential patterns and analyses will be conducted to determine if there is differential attrition by treatment arm, and if missingness is related to any of the covariates. If nonrandom missingness is of concern, a pattern-mixture, propensity score or related models will be applied so that bias effect can be assessed in sensitivity analyses.

This study also lacks a model assessing the costs and benefits of CHW HIV programs. While our prior RCT did not conduct a formal economic analysis, we estimated an annual cost of nearly $2,750 per intervention participant. While this intervention does not call for a formal cost analysis, data on health care utilization and hospitalizations are among the outcomes captured for this study. If linked to expenditure data, this information could be linked to expenditure data to develop a formal cost-effectiveness analysis for future studies.

## Conclusion

Results collected from INSTACARE will provide important information on the effectiveness of a clinic-based CHW intervention on improved HIV outcomes among diverse Black populations. The study highlights the innovative role of CHWs play in chronic care delivery, and our findings will further gauge the feasibility of this framework in the existing health care system.

**Spirit Figure 1.**
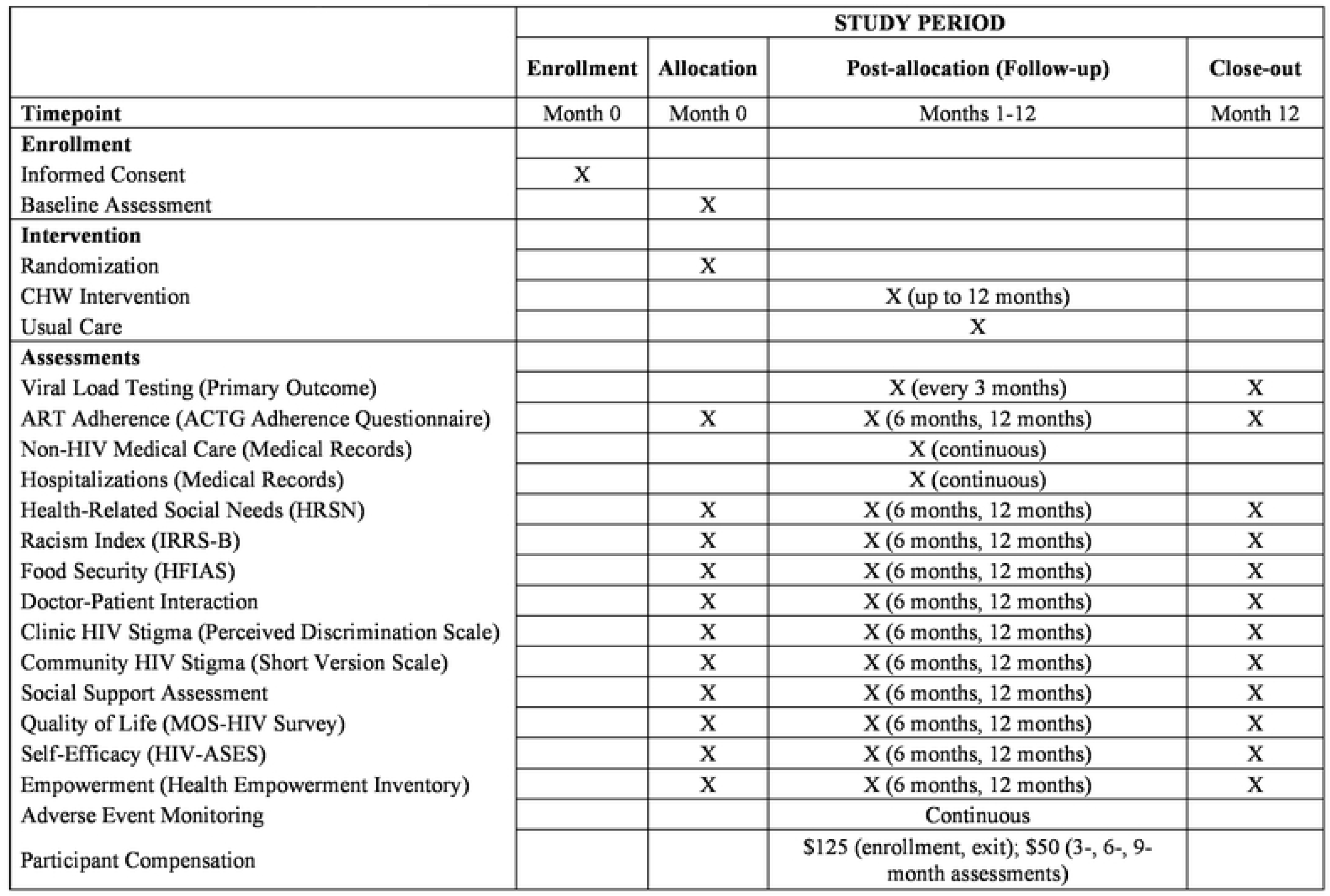
INSTACARE SPIRIT 2013 Figure

## Data Availability

No datasets were generated or analysed during the current study. All relevant data from this study will be made available upon study completion.

## SUPPORTING INFORMATION

S1 FIG. SPIRIT FIGURE

S2 TABLE. SPIRIT CHECKLIST

S1 FILE. INSTACARE PROTOCOL

## Trial registration

ClinicalTrials.gov NCT04663152

